# Cost-Effectiveness Analysis of the screening program for Diabetic Retinopathy at primary healthcare level in Patients with type 2 Diabetes Mellitus in Danang city, Vietnam

**DOI:** 10.64898/2026.01.02.26343334

**Authors:** Thi Hoai Vi Tran, Nguyen Quoc Dat, Nguyen Hoang Lan

## Abstract

**Objective:** This study is to evaluate the cost-effectiveness of an annual screening program for diabetic retinopathy (DR) at primary healthcare level among patients with type 2 diabetes mellitus (type 2 DM) from a societal perspective in Danang city, Vietnam.

**Methods:** A Markov model was developed to compare costs and effects of an annual screening program for DR using non-mydriatic fundus photography with the absence of the screening on a cohort of 23,951 patients with type 2 DM aged 40 years in Danang city from a societal perspective over a lifetime horizon of 40 years. The effect was estimated using quality-adjusted life years (QALY). Costs were measured in both Viet Nam Dong and US dollar. Both costs and effects were discounted annually at 3%. One-way and probabilistic sensitivity analyses were conducted to assess parameter uncertainty.

**Results:** The ICER for the DR screening program compared with the absence of screening was VND 86,631,252 per QALY (USD 3,630/QALY). In the one-way sensitivity analyses, a decrease in the screening participation rate, screening every two years, lower sensitivity of fundus photography and the poor treatment compliance rate, as well as an increase in costs of screening and treatment, which all led to higher ICER values. Increasing the incidence of DM did not change ICER value. Conversely, reducing treatment costs caused lower ICER. At a willingness-to-pay (WTP) threshold of VND 86,631,252 (USD 3,630) per QALY, the probability of the screening program being cost-effective was 92.3%. When the WTP increases at VND 95,294,377 (USD 3,993) or higher, the probability of cost-effectiveness approached 100%.

**Conclusion:** Diabetic retinopathy screening for patients with type 2 diabetes for aged 40 to 80 years is cost-effective in Danang city, Vietnam according to World Health Organization criteria.

## Introduction

Diabetic retinopathy (DR) is a microvascular complication of diabetes mellitus (DM), caused by damage to the blood vessels of the retina, leading to bleeding, plasma leakage and the growth of new abnormal blood vessels. These changes result in the formation of fibrous scar tissue, which can cause decreased vision or even vision loss (1). This is the most common complication of diabetic eye disease and the leading cause of blindness in patients, mainly affecting adults of working age (2), significant reduction in their quality of life (3–4) and create a considerable economic burden for patients, their families and the community. DR is a significant global public health and economic burden (5).

In Vietnam, there are currently more than 3.5 million people living with DM, most commonly adults aged 40 years and older (6). It is projected that the number of people with diabetes in Vietnam will rise to nearly 6.3 million by 2045 (7). In recent years, hospital-based studies reported that the prevalent rate of DR ranged between 20% and 58.7% among DM patients (8–10). Of these, patients with proliferative DR may accounted for up to 37% (11). With the increasing prevalence of diabetes, the number of people with DR is also expected to rise.

Early detection and timely management of DR contribute to the prevention of vision loss, improvement of quality of life in diabetic patients and reduction of societal healthcare costs (12), (2).

Screening programs for early detection of DR have been implemented in many countries worldwide. Various screening strategies have been adopted. In Taiwan, the average cost-utility ratio (ACUR) for annual screening was reported as NT$ 21,924, achieving cost effectiveness (13). Similarly, previous studies have shown that (14) systematic fundus photography compared to no screening provided an ICER of KRW 57,716,867 per QALY, achieving cost effectiveness as well. Besides, other study reported that community-based DR screening in patients aged ≥50 years was more cost-effective than non-screening program in China, with costs of USD 4,179 per QALY gained in rural areas and USD 3,812 per QALY gained in urban areas (15). Most these studies have illustrated that DR screening is cost-effective when compared to no screening.

Before July 15, 2025, Vietnam’s healthcare system is organized into three levels, consisting of primary healthcare level (PHC) (commune health stations, district health centers), city/provincial level and central level (tertiary hospitals). In this structure, eye care services are mainly delivered through district health centers (DHCs), which have mostly limited resources. Opportunistic screening for DR has been still mainly at city/provincial eye hospitals but the participation has been limited. Owing to the support of some international non-governmental organizations, systematic screening programs for DR were implemented in a few provinces and cities in the country. The mass screening programs for DR have not been deployed nationwide.

### Screening program for DR in Danang city

Danang is a city in central Vietnam with more than one million people and the average per capita income about VND 74,684,400 (3,129 USD) in 2023 (16). There were 23,951 patients with type 2 diabetes who were managed in the centers for disease control and prevention (CDC) of the city in 2023 (17).

With the support of Fred Hollows Foundation (FHF) under the project “Early detection of Diabetic Retinopathy in the Community”, community-based screening campaigns has been annually deployed between 2020 and 2023. The project had performed many activities including training in DR screening and referral skills for health staff in District health center, providing the non-mydriatic fundus photography to those facilities and publishing communication materials about DR. The DM patients were invited to examine eyes with visual acuity testing and fundus photography by trained technicians at district health centers (DHCs), then suspected DR cases were referred to Danang Eye Hospital for confirm diagnosis by ophthalmologists. By the end of 2023, the project had screened 8,133 DM patients among whom 232 patients with DR were detected (2.9%) (18). Those DR patients received appropriate treatment after diagnosis. Their DR treatment process was implemented according to the current care guidelines at the eye hospital as follows: patients diagnosed with mild to moderate non-proliferative DR were regularly followed-up based on the disease progression. Patients with proliferative DR or macular edema were treated laser or anti-VEGF (Vascular Endothelial Growth Factor) therapy. All services were funded by the FHF project and health insurance (18).

After closing the project, taking advantage of the project’s available resources, the department of Health of Danang city conducted a systematic screening program. Participants included 900 individuals with type 2 DM from 3 districts which represent three geographical areas of the city. The screening process for DR is similar to that of the FHF project. However, expenditure for the program was combined from different sources: program organization costs used the health department’s operating fund, costs for eye exam at DHCs, cost for confirmed diagnosis and treatment services come from health insurance and patients’ co-payments (19).

The aim of the study was to estimate the cost-effectiveness of the annual DR screening program at PHC from a societal perspective in Danang city. The findings of the study are expected to provide valuable evidence for national health program planning to reduce impact of eye complication of diabetes and to inform healthcare decision-making in Vietnam.

## Methods

A model in which both epidemiological and cost aspects for DR are included was designed to estimate the cost and effectiveness of a screening program for DR. Sensitivity analyses were also conducted to assess uncertainty of the modelling parameters

### Markov model structure

A Markov model was developed to simulate long-term costs and outcomes for the cohort of type 2 DM patients in Danang city. The model compared cost and effects of the annual screening program using non-mydriatic fundus photography with the same situation without such a program. The cohort included 23,951 type 2 DM aged 40 years free DR (17). The model estimated cost and effects over a period of 40 years with the assumption that the DM patients live to average life expectancy of the population in Danang city (20). Each year DM patients transit through one of six states following: 1) no DR, 2) mild non-proliferative DR (mild nPDR), 3) moderate non-proliferative DR (moderate nPDR), 4) severe non-proliferative DR (severe nPDR), 5) proliferative DR (PDR) and 6) blindness according to annual incidence of DR **(Fig 1)**.

**Fig 1.**
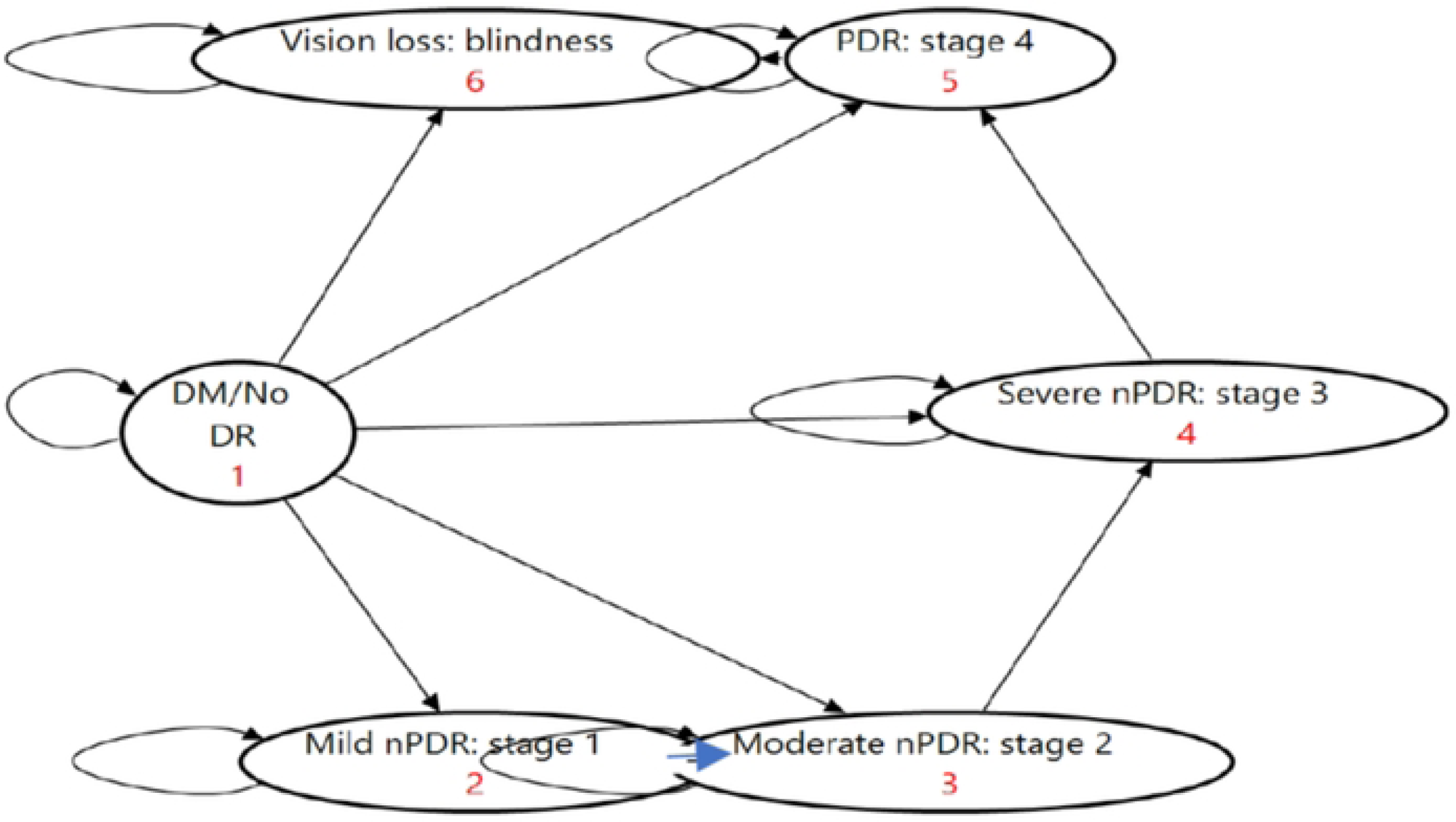
The structure of the Markov model.

For baseline model, we assumed that in screening arm, 100% of DM patients participated in screening program every year and all individuals who were diagnosed with DR received appropriate treatment until they became blind. In terms of both arms, DR incidence at each year was based on DR rates detected by the screening program (19) and those rates were stable over time. In each cycle (one year) patients could remain in their current state or transition to a more severe DR state, depending on transition probabilities **(Fig 2)**. The model also hypothesized that every patient had DR in only one eye to avoid overestimation of treatment costs and followed popular clinical care pathways.

**Fig 2.**
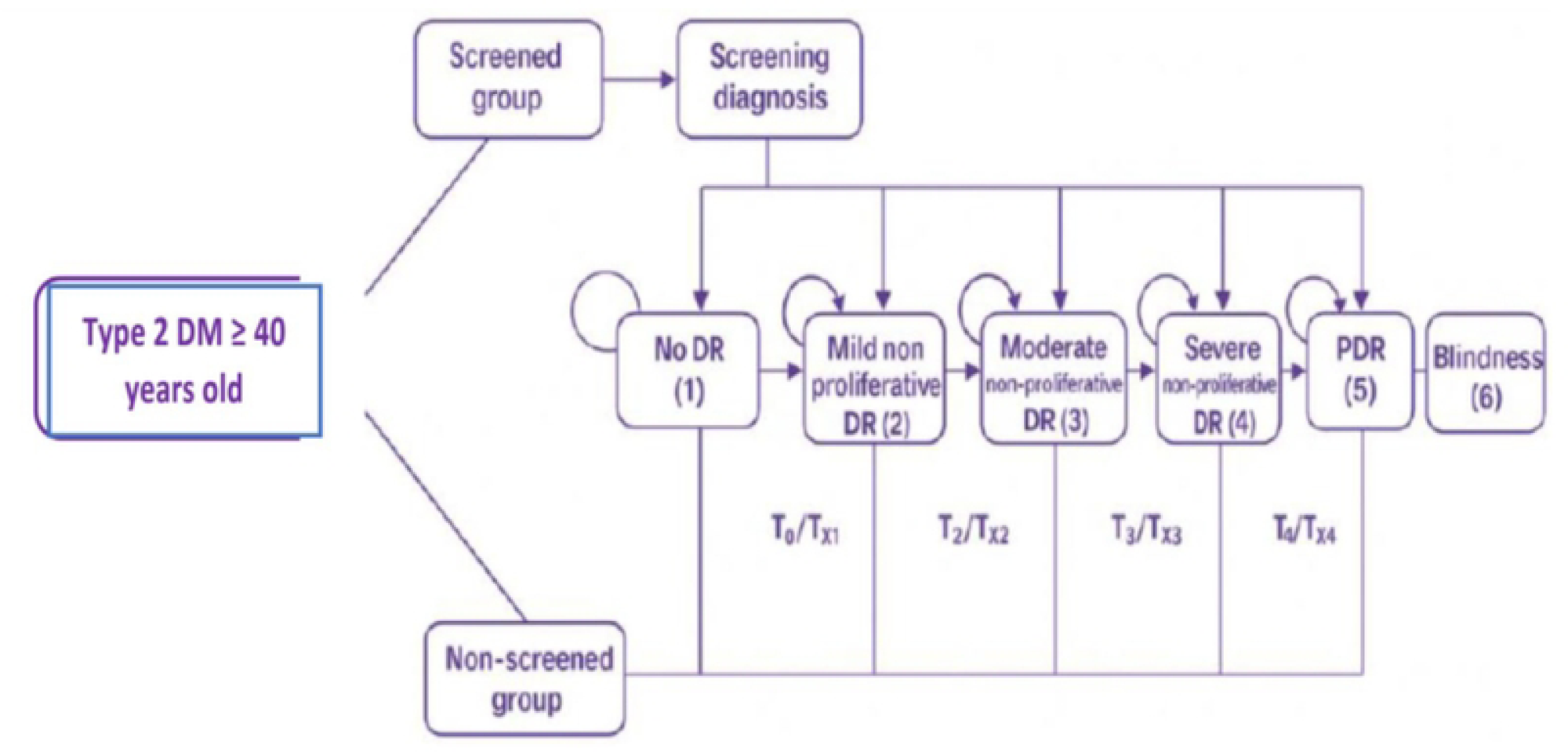
State transition diagram of Markov model.

### Perspective, time horizon, and discounting

The analysis was conducted from a societal perspective, including direct medical, direct non-medical, and indirect costs. In the absence-of-screening arm, only costs related to productivity losses due to vision impairment were considered, reflecting routine care without systematic screening. A lifetime horizon, defined as 40 years based on the average life expectancy of the cohort, was applied to capture long-term costs and health outcomes of DR screening. Both costs and QALYs were discounted annually at 3%, in line with World Health Organization recommendations.

### Input Parameters and Data Resources

#### DR Incidence

DR incidence in each cycle of the model was estimated based on rates of DR which was detected by screening program from 900 patients with type 2 diabetes in Danang city in 2023 (19). Stage- and age-specific DR incidence was calculated by age-specific incidence rates multiplied by the stage distribution of DR **(Table 1)**. It was assumed that these rates remained constant over time. The age-specific mortality rate and other diabetic complications were also considered the same in both arms, they were ignored in our model.

**Table 1.** DR incidence.

| Type | Parameter | Base value | 95% confidence interval (CI) | Source |
| --- | --- | --- | --- | --- |
| **Distribution of stage-specific DR** | No DR | 89.1% |  | (19) |
|  | Mild NPDR | 5.9% |  |  |
|  | Moderate NPDR | 3.1% |  |  |
|  | Severe NPDR | 1.0% |  |  |
|  | PDR | 0.9% |  |  |
|  | Blind | 0.0% |  |  |
| **Age-specific DR** | 40-44 | 0.0108 |  |  |
|  | 45-49 | 0.0441 |  |  |
|  | 50-54 | 0.0990 |  | (19) |
|  | 55-59 | 0.2088 |  |  |
|  | 60-64 | 0.2530 |  |  |
|  | 65-69 | 0.2196 |  |  |
|  | 70-74 | 0.1647 |  |  |
| Stage and Age-specific DR | Initial stage-specific DR * Age-specific DR |  |  | (19) |
| Natural transition probabilities | Mild NPDR to Moderate NPDR | 0.1796 | 0.0953-0.3127 | (21-25) |
|  | Moderate NPDR to Severe NPDR | 0.3404 | 0.1608-0.5816 |  |
|  | Severe NPDR to PDR | 0.4265 | 0.3648-0.4905 |  |
|  | PDR to Blind | 0.1739 | 0.0844-0.3245 |  |
| Treatment effectiveness | RR | 0.24 | - | (26-27) |
| Transition probabilities with treatment | Natural transition probabilities * 0.24 |  |  |  |

#### Transition probabilities between DR states

We applied two types of the transition probabilities between DR states in the model **(Table 1)**:

+ Natural transition probabilities were used in non-screening arm. The natural transition probabilities between DR stages were derived from a meta-analysis result (21–25).

+ Transition probabilities with treatment were used in screening arm, which was hypothesized that 100% of DR patients were adherence to the treatment. Transition probabilities incorporating treatment effectiveness were obtained from published studies (26–27), with a reported relative risk of 0.24.

#### Cost estimates

- Cost of DR diagnosis and treatment were estimated from perspective of society including patients and health insurance. Direct medical costs, direct non-medical costs and indirect costs (productivity losses associated with vision loss) were included in model in screening arm while only indirect costs were applied in no screening arm **(Table 2)**. Direct medical costs of diagnosis, treatment and follow-up were calculated based on price list of the regulated healthcare service in the Danang city’s eye hospital in 2023 (28). Direct non-medical (transportation cost) and proportion of income loss were collected by interviewing 207 DR patient of the city. Indirect costs were estimated on the basis of the city’s per capita income in 2023 and average annual of income loss rate for each stage of DR with the exception to patients aged 60 years and over who were retired.
- The screening costs included training costs, equipment procurement costs and fee of eye exam and fundus photography **(Table 2)**. Annual training cost and cost of equipment per person was calculated by depreciation formula as following:

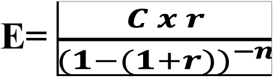

**Table 2.** Cost and Utilities using in the Markov model *(VND/USD)*

|  | No DR | Mild NPDR | Moderate NPDR | Severe NPDR | PDR | Blind |
| --- | --- | --- | --- | --- | --- | --- |
| **Screening costs/person** | **287,685 (12)** |  |  |  |  |  |
| *Training* | *5,391,669 (226)* |  |  |  |  |  |
| *Equipment* | *29,874,564 (1,251)* |  |  |  |  |  |
| *Fee of eye examination* | *192,600,000 (8,070)* |  |  |  |  |  |
| *Fee of fundus photography* | *31,050,000 (1,301)* |  |  |  |  |  |
| Total (900 patients) | 258,916,233 (10,848) |  |  |  |  |  |
| **Total diagnosis costs** | - | **385,196 (16.1)** | **472,132 (19.8)** | **1,695,341 (71)** | **1,656,191 (69.3)** | **448,800 (18.8)** |
| Direct medical cost | - | 303,800 (12.7) | 303,800 (12.7) | 1,167,600(48.9) | 1,167,600(48.9) | 303,800 (12.7) |
| Direct non-medical cost | - | 56,323 (2.3) | 99,545 (4.2) | 209,354 (8.8) | 172,676 (7.2) | 145,000 (6.1) |
| Indirect cost (the average income loss with participating in diagnosis via interviews) | - | 25,073 (1.1) | 68,787 (2.9) | 318,387 (13.3) | 315,915 (13.2) | 0 |
| **Total treatment costs (1st year)** | - | **574,123(24)** | **14,453,300(605.6)** | **49,770,999(2,085)** | **75,233,629(3,152.4)** | **80,020,151(3,352.8)** |
| Direct medical cost | - | 517,800 (21.7) | 11,274,302(472.4) | 31,614,506 (1,324.7) | 46,965,259 (1,967.9) | 5,190,751 (217.5) |
| Direct non-medical cost | - | 56,323 (2.3) | 199,090 (8.2) | 628,064 (26.3) | 777,042 (32.5) | 145,000 (6.1) |
| Indirect cost (productivity loss, <60 years old) | - | 0 | 2,979,908 (125) | 17,528,429 (734) | 27,491,328 (1,152) | 74,684,400 (3,129.3) |
| Indirect cost (≥60 years old) | - | - | - | - | - | - |
| **Total follow-up costs** | - | **360,123 (15)** | **8,728,504 (365)** | **39,728,755 (1,664)** | **60,263,862 (2,525)** | **74,684,400 (3,129.3)** |
| Direct medical cost | - | 303,800 (12.7) | 5,649,051 (236.7) | 21,780,004 (912.6) | 32,254,506 (1,351.5) | 0 |
| Direct non-medical cost | - | 56,323 (2.3) | 99,545 (4.2) | 420,322 (17.6) | 518,028 (21.7) | 0 |
| Indirect cost (productivity loss, <60 years old) | - | 0 | 2,979,908 (125) | 17,528,429 (734) | 27,491,328 (1,152) | 74,684,400 (3,129.3) |
| Indirect cost (≥60 years old) | - | - | - | - | - | - |
| **Utilities** | 0.95 | 0.86 | 0.80 | 0.71 | 0.64 | 0.31 |

In which:

E: annual cost
C: total expenditure
r: 3% (discount rate recommended by WHO) (29)
n: 10 years

i. Annual training costs were estimated under assumption that all healthcare staff trained by the program would continue working for 10 years. Total training expenditure from the FHF project (18) was allocated according to three districts among project seven districts.
ii. Equipment cost per person was calculated assuming a 10-year useful lifespan. Total equipment expenditure (fundus cameras) provided by the FHF project were depreciated and divided by the total number of screened participants in the FHF project (18).
iii. Fee of eye exam and fundus photography was based on regulated price list for DHCs in Danang city in 2023 (28).

All costs were adjusted to the year 2023 based on the Vietnamese Consumer Price Index (30) and measured in both Viet Nam Dong and US dollar ($) (exchange rate in 2023, 1 USD = 23,866 VND (31)).

#### Outcome estimates

The effectiveness of the screening program was expressed as quality-adjusted life years (QALYs). QALYs of each state of DR was received from a previous study in Danang city (32) **(Table 2)**.

### Cost- effectiveness analysis

An ICER (Incremental Cost-Effectiveness Ratio) was calculated by dividing the difference in total costs by the difference in QALYs gained between the two arms.

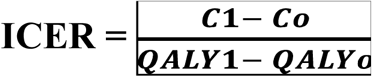

In which:

C_1_: Total cost of the screening program.

C_0_: Total cost of the no-screening program.

QALY_1_ : Total QALYs gained from the screening program.

QALY_0_: Total QALYs of the no-screening program.

Based on the recommendation of World Health Organization (WHO) (33) for cost-effectiveness of an intervention, we used income per capita of Danang city to assess cost-effectiveness of the screening program in the study.

1. Less than income per capita: highly cost-effective.
2. Between one and three times income per capita: cost-effective.
3. More than three times income per capita: not cost-effective.

### Sensitivity analyses

Both one-way sensitivity analysis and probabilistic sensitivity analysis were conducted.

#### One-way sensitivity analysis

A series of one-way sensitivity analysis was performed on the following parameter: the increase in prevalence of DM, cost of screening, treatment/follow-up and the reduce in sensitivity of the screening device, screening participation rate, treatment/follow-up adherence rate, screening frequency and cost of treatment/ follow-up **(Table 3)**.

**Table 3.** One-Way Sensitivity Analysis of Key Parameters for DR Screening.

| Sensitivity Analysis<br>Parameter | Base Case | Analysis Value | Source |
| --- | --- | --- | --- |
| **1. Epidemiological parameters** |  |  |  |
| Prevalence of DM | 7.4% | 10.8% (46% increase by<br>2045) | (6), (34) |
| **2. Screening device (non-mydratic fundus camera)** |  |  |  |
| Sensitivity | 97.0% (1) | Reduce 82.13% | (35-37) |
| **3. Parameters related to screening participation and treatment adherence** |  |  |  |
| Screening participation<br>rate | 100% | 70% | Assumption |
| Treatment/follow-up<br>adherence rate | 100% | 80% | Assumption |
| Screening interval | Annual | Biennial | Assumption |
| **4. Cost parameters** |  |  |  |
| Screening costs | VND 287,685 (\$12) | +20% | Assumption |
| Treatment/follow-up<br>costs (health service<br>costs) | Cost in 2023 | +20% | Assumption |
|  |  | -20% | Assumption |

#### Probabilistic sensitivity analysis

Uncertainty parameters were assigned suitable probability distributions as recommended by Briggs et al. (38) in the probabilistic sensitivity analysis **(Table 4)**. Monte Carlo simulation using with 2000 interactions to assess the impact of uncertainty about the modeling parameters on the ICER estimates. The result of the PSA was displayed by ICER scatter plot in a cost-effectiveness plane. A cost-effectiveness acceptability curve, which shows the probability of the annual DR screening being cost-effectiveness over a range of willingness to pay thresholds was also created from this finding. TreeAge Pro 2025 software was used to construct and run the model.

**Table 4.** Distribution for uncertainty parameters.

| Uncertainty parameters | Distribution |
| --- | --- |
| Initial stage-specific DR | Beta |
| Age-specific DR | Beta |
| Utilities | Beta |
| Transition probabilities | Beta |
| Cost | Gamma |

## Ethics Approval

Ethics approval for this study was issued by the Research Ethics Board of Hue University of Medicine and Pharmacy (Approval No. H2022/521, dated December 30, 2022).

## Reporting standards

This study is reported in accordance with the Consolidated Health Economic Evaluation Reporting Standards (CHEERS 2022).

## Results

### Baseline Results

Base-case results are reported in Table 5. Implementation of the annual screening program for DR in type 2 DM patients starting at age 40 years and continuing for 40 years increased the number of quality-adjusted life gained from 30.80 to 35.16 years per person. The average cost of the program on the basis of society’s perspective was higher VND 72,270,510 ($3,028) compared to VND 450,396,429 ($18,871) per person with the absence of the program. The estimation of incremental cost-effectiveness ratio was VND 86,631,252 ($3,630) per QALY gained.

**Table 5.**
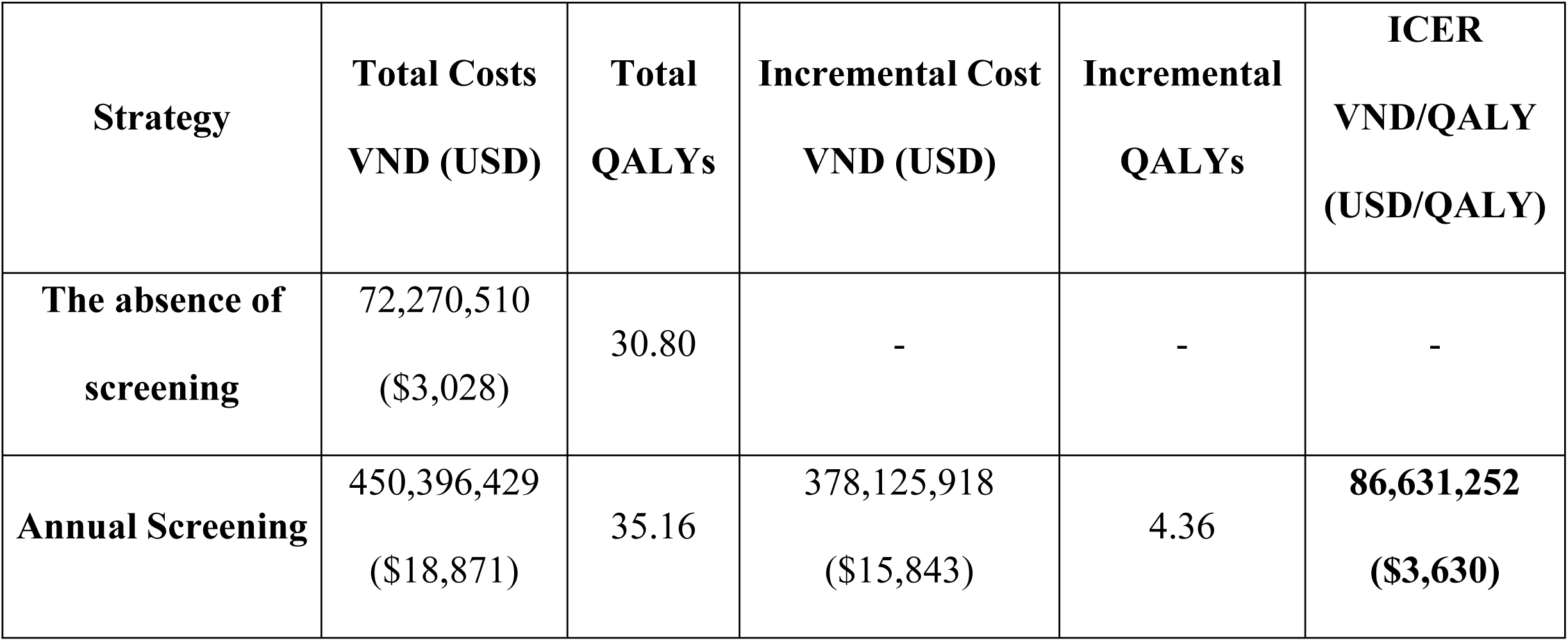
Incremental cost-effectiveness in base-case analysis.

### Sensitivity Analysis Findings

#### One-Way Sensitivity Analyses

The results of the one-way sensitivity analysis assessing the impact of various factors on the cost-effectiveness of the screening program were presented in a tornado diagram **(Fig 3)**. The decrease in the participation rate from 100% to 70% increased the highest, ICER value to VND 387 million ($16,215) per QALY. Followed by, reducing screening frequency to once every two years raised the ICER to VND 156.5 million ($6,557) per QALY. Then a rise in treatment costs of DR by 20% increased ICER value 105 million ($4,399) per QALY. Other factors including reducing the adherence to DR treatment therapy after screening by 20%, lowering the sensitivity of non-mydriatic handheld fundus cameras by 15% (from 97% to 82%) and raising cost of screening program by 20% slightly increased the ICER to VND 87,705,425 ($3.674), 87,307,706 ($3,658), 87,086,699 ($3,649) per QALY, respectively. With the increase in DM prevalence from 7.4% to 10.8%, ICER value remains unchanged.

**Fig 3.**
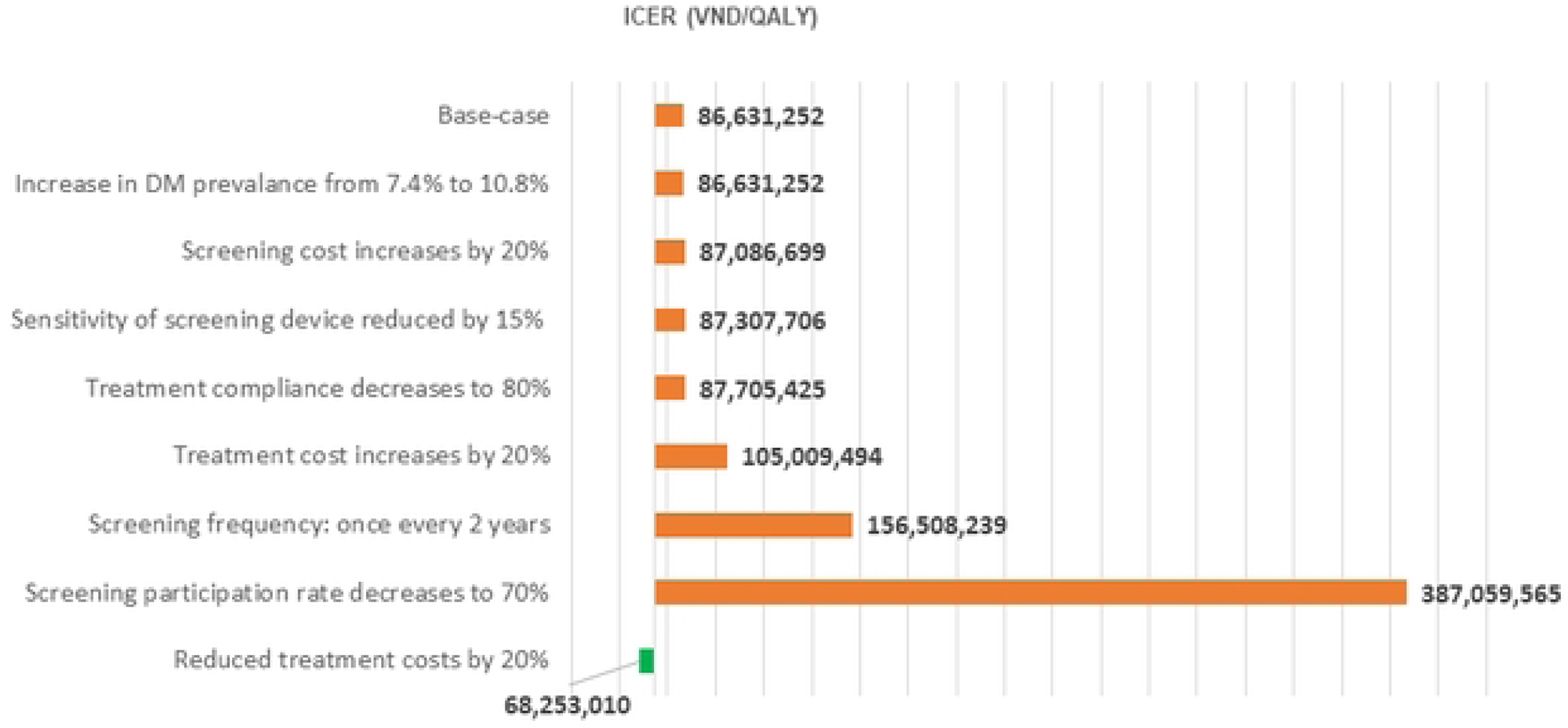
Tornado Diagram presenting results of one-way sensitivity analysis.

On the other hand, the decrease in the treatment costs reduced significantly ICER to VND 68,253,010 ($2,859)/QALY.

### Probabilistic Sensitivity Analyses

#### Cost-effectiveness plane (CE plane)

Fig 4 shows the difference in QALYs per person against the difference in cost per person. Each dot on the C-E plane represents a single output from probabilistic model. All observations were in the northeast quadrant of the C-E plane, thereby indicating that the screening program increased costs and improved effectiveness. In addition, the points form a tight oval-shaped clusters, suggesting that the ICER results are stable. ICER ranged from VND 75.870.382 ($3,179) to VND 92.078.166 ($3,858). The joint density did not yield any incremental cost below approximately VND 370,000,000 ($15,503) in the C-E plane. This implies that obtaining the additional health benefits of the screening program requires at least an additional cost of about VND 370,000,000 ($15,503) per person.

**Fig 4.**
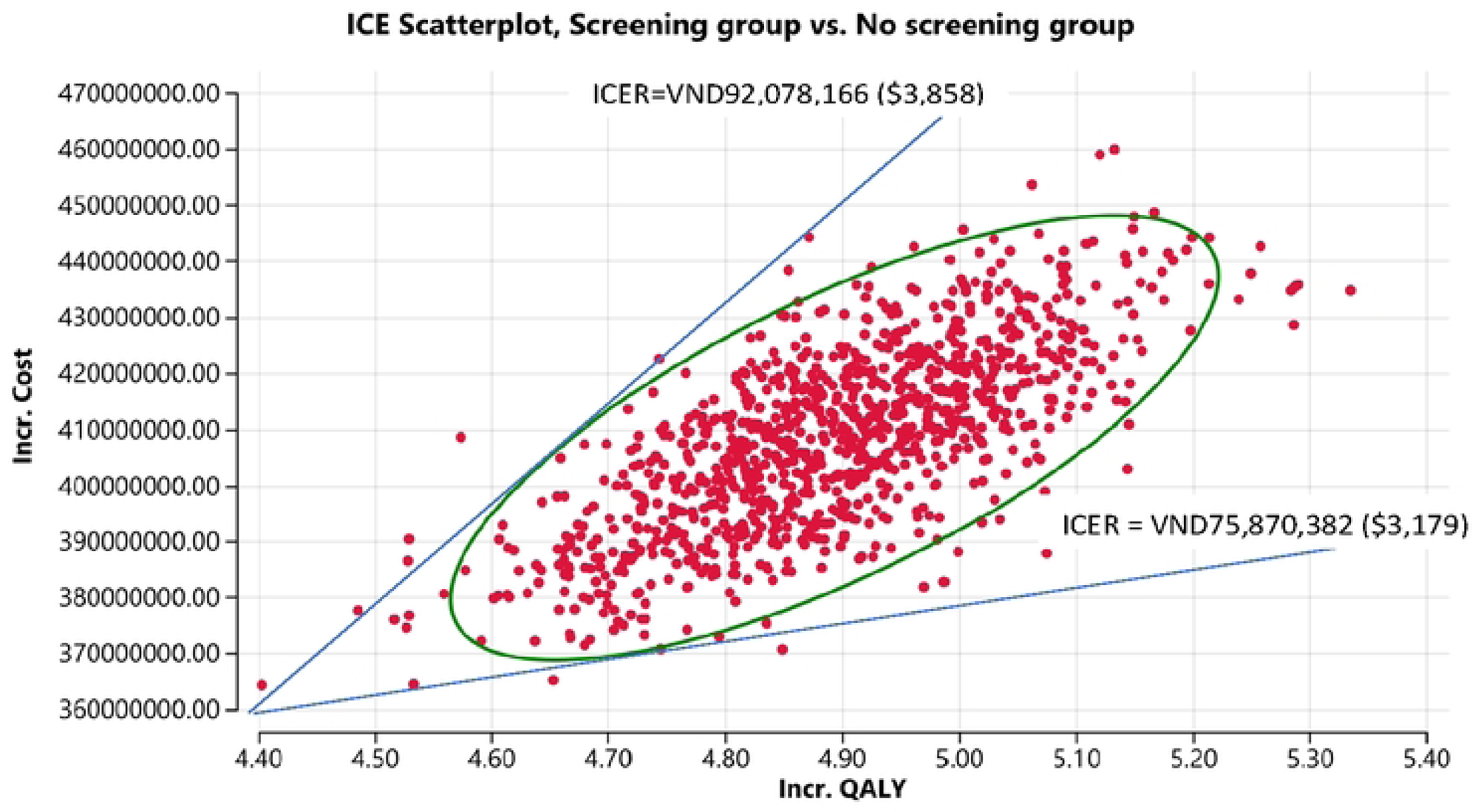
ICER scatter plot (C-E plane)

### Cost-effectiveness acceptability curve (CEAC)

The CEAC for the DR screening program is shown in Fig 5. The CEAC shows the probability that the screening program is cost-effective for different monetary values placed on QALYs gained. A greater willingness to pay (WTP) threshold for QALYs gained increases the probability that the screening program is more cost-effective. At a WTP threshold of VND 86,631,252 ($3,630) per QALY, the probability of the screening program being cost-effective is 92.3%. When the WTP increases beyond this threshold, reaches VND 95,294,377 ($3,993) or higher, the probability of cost-effectiveness approaches 100%.

**Fig 5.**
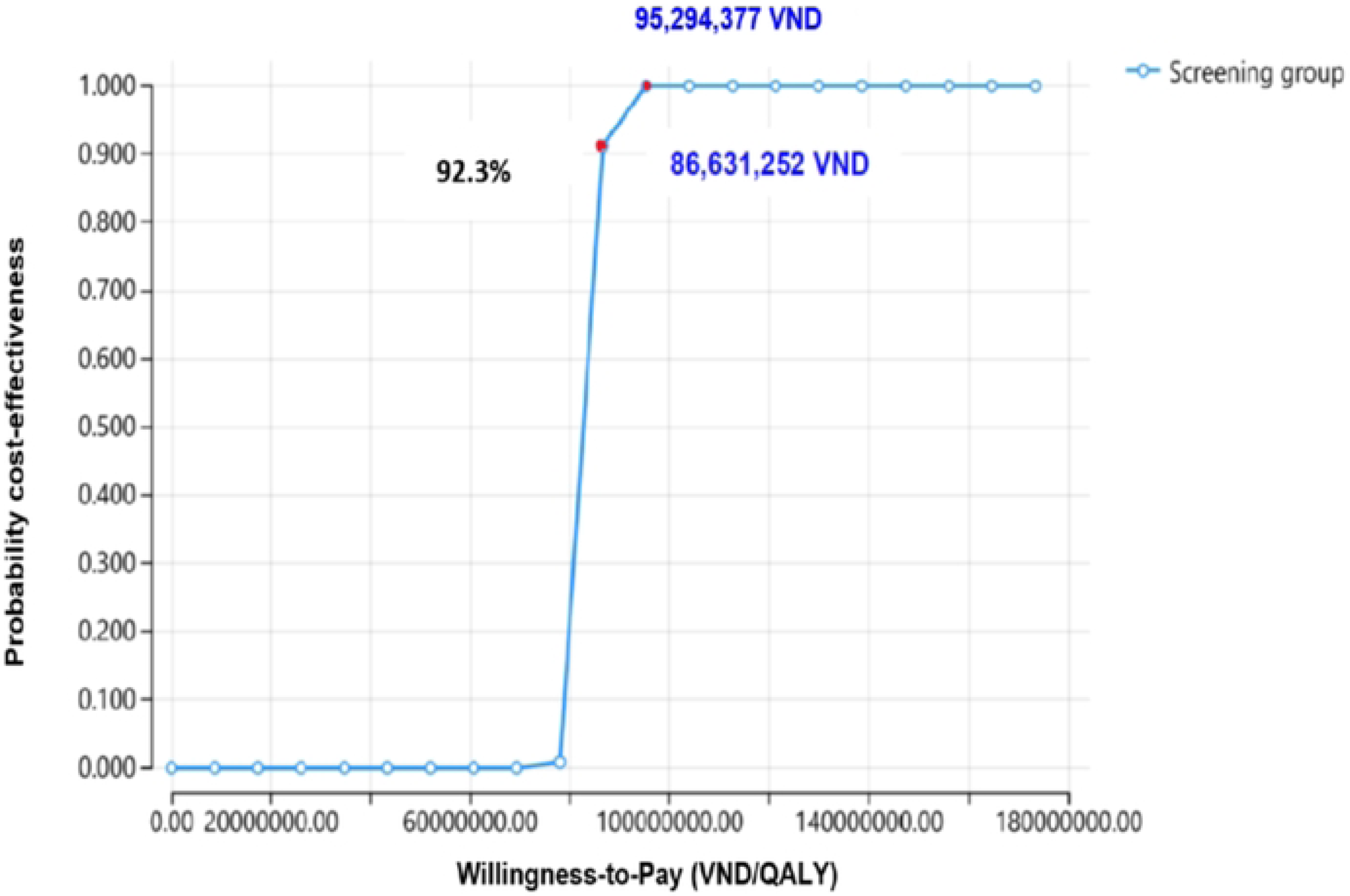
Cost-effectiveness acceptability curve for DR screening in Danang city.

## Discussion

### Base-Case Results

The baseline analysis has shown that the implementation of an annual screening for DR for type 2 DM patients from 40 years old in Danang city would enhance quality-adjusted life years gained, however the increase in the number of DR detected and treatment earlier increased cost incurred per patients by VND 378,125,918 ($15,843). Consequently, the incremental cost-effectiveness ratio (ICER) of the screening program was estimated at VND 86,631,252 ($3,630) per QALYs. Compared with average income per capita in Danang city and GDP Vietnam in 2023 (VND 101,900,000 ($4,269) (16), the screening program for DR in DM type 2 patients at PHC level was considered as cost-effective according to criteria of WHO (33). However, achieving these benefits requires a substantial investment of at least VND 378 million ($15,843) per person. This supports the program’s economic value but highlights the need to consider budget impact in decision-making. The result is lower than previous studies in the world using the similar screening strategy (14) (15). Higher unit cost increased ICER value of the studies. Screening cost with fundus photography and refraction tests in a study of Korea was $13.7 per person (14), comparing with $12 in our study. In China, the cost of medication (anti-VEGF) per person was $1,741 comparing with mean from $12.7 to $1,961 depend on stage-specific of DR in our study, and its non-medical direct cost was also higher than ours ($63 vs. $32) (15). The screening program in Danang city was provided at primary healthcare facilities to save expenses for travel and accommodation. Currently, Vietnam has re-structured health care system nationwide, in which DHCs become Regional health Centers with larger coverage, systematic DR screening in DHCs will more advantageous.

Monte Carlo simulations confirm the economic effect of the screening program. Table 5 shows all results in the northeast quadrant, meaning the screening program increases both costs and effectiveness. The ICER is stable, ranging from VND 75.8 to VND 92 million ($3,179 - $3,858) per QALY, all below three times the per capita income and below one GDP per capita of Vietnam, indicating that the screening program for DR in Danang city in especially and in Vietnam in general is cost-effectiveness. Further analysis on the cost-effectiveness acceptability curve **(Fig 5)** reported that an increase in the WTP threshold from VND 86,631,252 ($3,630) to VND 95,294,377 ($3,993), less than three times the per capita income in Danang city in 2023 and below one GDP per capita of Vietnam, would raise the cost-effectiveness probability of the screening program from 92.3% to 100%. The PSA showed that the probability of the intervention being cost-effective increased with a greater value of willingness to pay. This indicates that the screening program remains highly cost-effective under higher investment budget. The finding supports policy of public funding for systematic DR screening, especially as Vietnam considers providing free or subsidized screening services for chronic diseases. However, it is necessary to expand scale of the study and conduct a budget impact analysis of the screening program for DR to confirm economic benefits of the program nationwide.

A one-way sensitivity analysis was conducted to identify which individual parameters had the greatest impact on the ICER estimates. In this study, screening participation rates had the strongest influence on the estimated ICER’s. When the participation rate drops from 100% to 70%, there is a significant increase in the ICER to VND 387 million ($16,215) per QALY that is more than three times the per capita income in Danang city and more than three times GDP per capita of Vietnam, indicating that the screening program is no longer cost-effective. The explanation is that DR detection is late resulting in a higher proportion of patients progressing to severe stages of DR, at which treatment becomes more expensive. Similar conclusions were reported previously (14), the cost-utility analysis in Korea identified screening compliance as a key sensitivity factor. The study emphasized that if the compliance rate fell more than 10%, the assessment of the ICER is significantly affected. The policy implication is that attendance rates must be discussed alongside systematic screening strategies (14). Lack of awareness of DR in patients with DM has been considered an important risk factor for the poor attendance to screening programs designed for the purpose of disease management (39–40). Additionally, in Vietnam, health insurance fund has not cover most NCDs preventive services including screening (41–42). Therefore, beside a systematic screening program for DR should be developed, package of health insurance should cover fees of screening services and counselling for individuals suffering diabetes in Vietnam to ensure detection of DR at earlier stages.

Screening frequency is also a key factor influencing the cost-effectiveness in the study. Our sensitivity analysis showed that extending the interval to once every two years nearly doubled the ICER (VND 156,508,239 ($6,557) per QALY), meaning that biennial screening remains marginally cost-effective. The higher costs of the program is associated with delayed detection. Evidence from Kinmen, Taiwan compared with the control group (no screening), the annual screening program, as well as all longer intervals (biennial, 3-year, 4-year, 5-year), was found both more effective and less costly (13). According World Health Organization reports from 48 countries (43) show that many used annual screening intervals, while others screen every two years or even every 3-5 years. The choice of interval varies depending on resources, infrastructure, findings of screening and the risk stratification of T2DM patient groups (43). Nonetheless, the findings are largely consistent in highlighting that shorter screening intervals, whether annually or every two years to ensure timely detection and treatment of DR, while optimizing the allocation of healthcare resources (13), (43–44).

Increasing in costs of screening and treatment are also contributed to raise the ICER estimates, however, the ICER remains acceptable cost-effective thresholds (less than three times the per capita income of Danang city and GDP per capita of Vietnam in 2023). The incremental costs in the annual screening arm are mainly due to the implementation of screening and the treatment of detected cases. In particular, treatment costs with anti-VEGF therapies (45) account for a large proportion of total expenses in DR management and strongly influence cost-effectiveness outcomes. In Vietnam, including Danang city, treatment choices are strongly influenced by health-insurance reimbursement. Lucentis (Ranibizumab) is widely used because it is reimbursed - based on the health insurance copayment rate, despite its high cost. Avastin (Bevacizumab) is an alternative and cheaper medication but has been shown to have efficacy comparable to Lucentis (46), it is prescribed less frequently because it is only partially reimbursed (50%). High cost of anti-VEGF makes therapy a major driver of overall DR treatment costs. According to a study in Vietnam, cost of anti-VEGF accounted for more than 90% of the total expenses in DR treatment (45). Our results suggest that reducing the treatment costs by 20% would lower the ICER to below one time the per capita income, indicating that the intervention is highly cost-effective (VND 68,253,010 ($2,859) per QALY). Enhancing insurance reimbursement for Avastin would not only increase economic effectiveness of the DR screening program, but also reduce the financial burden on both patients and health insurance system. Importantly, lowering out-of-pocket costs is likely to improve patient adherence to recommended treatment, thereby enhancing clinical outcomes and maximizing the overall benefits of early DR detection. Besides treatment expenses, screening costs also contribute to the incremental costs in the screening arm. Although the annual screening cost per patient is relatively low, it becomes substantial when applied to a large DM population. As suggestion earlier, a budget impact analysis is necessary for decision makers to face the increase in expenditure on the program.

Treatment compliance was also a key factor that directly impacts the cost-effectiveness and sustainability of DR screening programs. Reducing treatment compliance rate by 20% slightly increased the ICER to VND 87,705,425 ($3.674). In line with the findings from our study, the Taiwan study found that the economic benefits (QALYs gained) estimated are fundamentally based on the extra time patients gain to seek treatment. Strong economic gains associated with early detection will be lost if patients do not comply with follow-up and treatment (13). DR is a chronic condition that requires ongoing management with regular follow-up appointment to monitor the response to treatment, detect any recurrence or progression of the disease early. A study in Vietnam found that high cost of treatment, transportation cost due to distance to eye hospitals and side effects of the treatment and side-effect of treatment which were key barriers to adherence of the treatment among DR patients (42). Reducing out-of-pocket expenses and improving effect of the treatment can be essential measures to maintain treatment adherence and preserve the long-term cost-effectiveness from systematic DR screening program.

The reduction of sensitivity of the fundus photography by 15% (82.13%) also slightly increased the ICER. An acceptable sensitivity threshold of 83-85% for handheld fundus cameras in eye disease screening, particularly for DR, has been reported previously (47). A decrease in sensitivity means a reduced ability to detect individuals who truly have the disease and an increase in false-negative cases (48), which not only result in increase in costs of treatment, but also prevent to detect sight-threatening DR. Investing in training health staff in fundus photography and DR lesion grading and appropriate diagnostic equipment at PHC level should be solutions to achieve optimum of the screening program for DR.

Although an increase in the prevalence of DM is not alter the ICER value, it would substantially raise the overall budget required to sustain the screening program due to the larger number of individuals needing to be screened. This highlights the importance of strengthening diabetes prevention and control strategies, such as promoting healthy lifestyles, improving early diagnosis of DM and enhancing community-based prevention programs to slow the growth of new DM cases and then reduce future screening burdens.

## Limitations and Considerations

This is the first study on cost-effectiveness of a screening program for DR in Vietnam. Although the study provides valuable insights, several limitations should be acknowledged. Firstly, data for several parameters were unavailable in Danang-Vietnam, some estimates were adopted from studies conducted in other countries. Secondly, age-specific mortality and other related complications of DM were assumed to be the same in both arms and ignored in the model, which may affect the accuracy of long-term ICER estimates. Thirdly, the base model assumed 100% annual screening participation and 100% treatment adherence, an idealized assumption that is unlikely to be achieved in real-life, however the one-way sensitivity analysis showed their impact on ICER values. Fourthly, the estimated treatment costs for DR patient with one eye and followed popular clinical care pathways could lead to underestimation of the cost of DR management. Lastly, the screening was conducted only in three representative regions with a limited number of participants and cost data as well as quality of life of DR patients were collected in Danang only, therefore the findings may lack national representativeness, thus, expanding the research to a nationwide scale is necessary to ensure more comprehensive evidence for national health policy planning.

## Conclusion

Annual DR screening using non-mydriatic fundus photography for patients with type 2 DM at primary health care is cost-effective from a societal perspective in Danang city according to World Health Organization criteria. The findings suggest expanding DR screening strategies using a risk-based approach at PHCs level in Vietnam through improving reimbursement of health insurance fund to ensure affordability, adherence and cost-effectiveness of the DR screening program.

## Data Availability

All relevant data are included within the article. Model inputs were obtained from published literature and publicly available data sources, which are cited in the article

## Acknowledgments

The authors would like to thank the Danang Department of Health and participating healthcare facilities for their support in data collection. We also acknowledge all individuals who contributed to this study.

